# Safety, Tolerability, and Immunogenicity of PIKA-Adjuvanted Recombinant SARS-CoV-2 Spike (S) Protein Subunit Vaccine in Healthy Adults: Interim results of an open-label and randomised Phase 1 clinical trial

**DOI:** 10.1101/2022.11.20.22282565

**Authors:** Yuan Liu, Lai Hock Tan, Nan Zhang, Yi Zhang, Zenaida Reynoso Mojares

## Abstract

**Background:** This COVID-19 pandemic has caused unprecedented morbidity, mortality, and global economic instability. Several approved vaccines demonstrated to be effective prevention against COVID-19. We aimed to evaluate the safety and immunogenicity of the PIKA-adjuvanted recombinant SARS-C0V-2 Spike (S) protein subunit vaccine in adults as a primary immunization and as a booster dose against SARS-C0V-2 infection.

**Methods:** This was a Phase I, open label, dose-escalation study of 3 dose levels of the SARS-CoV-2 spike antigen administered intramuscularly in combination with a fixed dosage of PIKA adjuvant vaccine to evaluate the safety, tolerability, and immunogenicity of PIKA COVID-19 vaccine candidate in healthy adults. The study planned to have 3 arms: Arm A included subjects who had never received any Covid 19 vaccination or have had Covid 19 infection for > 6 months prior to enrolment, Arm B1 included subjects who had completed their primary series of Covid 19 vaccination with an inactivated Covid 19 vaccine and Arm B2 which included subjects whose primary series was completed with mRNA Covid 19 vaccine. The primary safety outcome was adverse events and safety laboratory parameters, and the secondary immunogenicity outcome was neutralizing antibody geometric mean titers and seroconversion rates against the wild type virus, Delta and Omicron variants. This trial is registered with ClinicalTrials.gov, number NCT05305300.

**Findings:** This interim analysis report presented the results of Arm A and Arm B1 who completed Day 35 for 2 doses in Arm A and Day 28 for a single booster dose in Arm B1.

*Safety results:* Arm A: 60% of participants reported mainly solicited AEs after first and second vaccine. Most of those were local (mainly pain/tenderness) with few systemic (mainly fever and headaches). The majority of participants reported unsolicited events after vaccine which were mainly investigations in hematology/hepatobiliary/Renal or Urine tract infection urine analysis. At least 80% of the participants reported mild AEs. There were 4 SAEs that were mild and were resolved. Also there were 2 medically attended AEs. Arm B1: Less than 50% of the participants reported solicited adverse events which were mainly local (pain and tenderness) and were mild. Also, less than half of the participants reported unsolicited events which were mainly investigations in hematology/hepatobiliary/Renal or Urine tract infection urine analysis. There were no SAE and Medically attended AEs reported.

*Immunogenicity results:* Arm A: The neutralizing antibody GMTs at day 35 were substantially higher than those at baseline for all dose groups and all variants. Seroconversion rates at 35 days ranged between 85.7% and 92.9% for 5μg dose group, 92.9% and 100% for the 10μg dose group and between 70% and 80% for the high dose group. Arm B1: Similar to Arm A, neutralizing antibody GMTs at day 28 were substantially higher than those at baseline for all dose groups and all variants. Seroconversion rates at 28 days ranged between 92.9% and 100% for 5μg dose group, 80% and 100% for the 10μg dose group and between 50% and 64.3% for the high dose group.

**Conclusion:** The findings demonstrated that the PIKA Covid 19 vaccine is safe, well tolerated, immunogenic and can be used as a primary vaccination or as a booster dose in participants who had completed an inactivated Covid 19 vaccination series. A comparison of the immune responses presented in this interim analysis showed that geometric mean titer (GMTs) of neutralizing antibody against wild type of SARS-CoV-2 virus, Delta and Omicron of the 5μg group was higher than the 10 μg and 20 μg, therefore the 5μg was selected as the recommended dose for the Phase II and III clinical development of the PIKA Covid 19 vaccine.

## Introduction

Severe acute respiratory syndrome coronavirus 2 (SARS-CoV-2), as the causative agent of coronavirus disease 2019, also known as COVID-19, as a beta-coronavirus which belongs to the family of Coronaviridae and is currently responsible for the third human coronavirus outbreak in the past 20 years after SARS (now often referred to SARS-CoV-1) in 2003 and MERS (Middle East respiratory syndrome) in 2012^1^. This COVID-19 pandemic has caused unprecedented morbidity, mortality, and global economic instability. Several approved vaccines including mRNA and adenovirus-vectored vaccines demonstrated to be effective against COVID-19.

Due to its involvement in the viral entry, the S protein is a major target for current vaccine development against SARS-CoV-2. The majority of the vaccine candidates under development currently use full length or truncated S protein^4-5^. Preliminary efficacy data from several Phase 3 trials showed promising results that spike antigen-based vaccine conferred high level protection against COVID-19, severe COVID-19 in particular. Emerging evidence indicates Spike protein-based vaccine also provide protection against variants. Therefore, we explored the recombinant SARS-CoV-2 protein as a potential vaccine candidate for the prevention of COVID-19 or SARS-CoV-2 infection.

The investigational PIKA COVID-19 vaccine is a combination of the SARS-CoV-2 spike (S) subunit protein and the PIKA adjuvant. The final formulation of PIKA COVID-19 vaccine is in a clear colourless solution form. The volume for a standard vial is 1ml. Three doses of PIKA COVID-19 vaccine were evaluated in this study to determine the selection of the dose and dosing regimens for future studies.

PIKA adjuvant has been developed by Yisheng Biopharma and is a chemically synthesized double-stranded RNA analogue, which is comprised of double-stranded polyinosinic acid-polycytidylic acid, kanamycin monosulfate and calcium chloride. It is an antagonist of toll-like receptor 3 (TLR3), a key pathogen associated molecular pattern receptor for recognizing dsRNA virus infections, that activates the innate immune system^6.7.8^. PIKA is an excellent adjuvant because it has been found to: 1) to interact specifically with TLR3 in a dose-dependent manner; 2) to promote the activation and maturation of dendritic cells and upregulate the co-stimulatory molecules, such as CD80, CD86, HLA-DR, CD83, and MHC-Class I on dendritic cells; 3) to induce the activation and proliferation of both B and NK cells; 4) to elicit both Th1 and Th2 immune responses. As an adjuvant, PIKA specifically interacts with TLR3 to activate dendritic cells, antigen-presenting cells that effectively activate B cells and T cells, thereby inducing an effective immune response^8,9^.

## Methods

### Study Design

We did a single-centre, open-label, randomised phase I trial in El Kuwait Hospital (El Baraha Hospital) in United Arab Emirates (UAE). The study was designed by Yisheng Biopharma (Sponsor) and conducted under the supervision of the principal investigator, at the Al Kuwait Hospital. Investigators were selected based upon their medical expertise, their willingness to conduct the study according to GCP requirements, and their ability to enroll eligible subjects.

Prior to the start of the study, the study protocol, protocol amendments, the informed consent form (ICF) and study related documents were approved by the Independent Ethnics Committee (IEC). The study was carried out in compliance with the protocol, in accordance with the International Council for Harmonization Technical Requirements for Registration of Pharmaceuticals for Human Use (ICH) harmonized tripartite guideline for Good Clinical Practice (GCP) and MOHAP (Ministry of Health and Prevention UAE) Research Ethics Committee.

Prior to subject participation in the study, written informed consent was obtained from each subject according to the regulatory requirements of UAE and each signatory dated their signature. The ICF provided subjects with information regarding the purpose, procedures, requirements, and restrictions of the study, along with any known risks and potential benefits associated with the investigational product (IP). The patient consent form and all other documents provided to the subject were translated from English into the appropriate native language of the individual concerned.

The study comprised of two arms. Arm A included subjects who have not received COVID-19 vaccine or with a history of COVID-19 infection for not less than 6 months prior to study participation and Arm B included subjects who received either an inactivated or mRNA Covid 19 vaccine as primary series.

In arm A, there were 3 dose groups, 1 to 3 with escalating antigen dose with PIKA adjuvant in sequential cohorts. A total of 45 subjects wereenrolled in Arm A. Fifteen eligible subjects in each dose group (5ug, 10ug and 20ug) received the study vaccine on Study Days 0 and 7 via single intramuscular injection in alternative deltoid muscles. Subjects were sequentially assigned to a dose group based on the timing of completion of screening. Enrollment of each dose cohort started with 3 sentinel subjects. The remaining subjects were enrolled at least 48 hours later after the 3 sentinel subjects completed the first dose administration with no safety concerns revealed. The 3 first subjects were followed up by phone call every 24 hours during the first 48 hours after vaccine administration. The medical monitor and principal investigator reviewed the safety experience of all subjects enrolled in Group 1 through Study Day 7. When no pausing rules had been met, Group 2 subjects were enrolled following the enrollment practice in Group 1. Accordingly, Group 3 subjects were enrolled when no pausing rules were met after all subjects in Group 2 completed Day 7 visit. The enrollment of Group 3 also followed the same enrollment practice as Groups 1 and 2.

In arm B, there were 3 dose groups (5ug, 10ug and 20ug), 1 to 3 with escalating antigen dose with PIKA adjuvant in sequential cohorts. A total of 90 subjects were enrolled in Arm B which were divided into Arm B1 (45 subjects) and Arm B2 (45 subjects). Arm B1 enrolled subjects who received COVID 19 Inactivated vaccines as primary series and was comprised of 15 subjects per dose (45 subjects). Arm B2 enrolled subjects who received COVID 19 mRNA vaccines as primary series and was comprised of 15 subjects per dose (45 subjects). Subjects in Group 1-3 of Arm B1 and Arm B2 received one booster dose of PIKA COVID-19 vaccine via IM administration on Day 0.

In Arms B1 and B2, fifteen eligible subjects in each dose group received the study vaccine on Study Days 0 via intramuscular injection in alternative deltoid muscles. Subjects were sequentially assigned to a dose group based on the timing of completion of screening. Enrollment of each dose cohort started with 3 sentinel subjects. The remaining subjects were enrolled at least 48 hours later after the 3 sentinel subjects completed the first dose administration with no safety concerns revealed. The 3 first subjects were followed up by phone call every 24 hours during the first 48 hours after vaccine administration. The medical monitor and principal investigator reviewed the safety experience of all subjects enrolled in Group 1. When no pausing rules had been met, Group 2 was enrolled following the enrollment practice in Group 1. Accordingly, Group 3 was also enrolled when no pausing rules were met after all subjects in Group 2 completed Day 7 visit. The enrollment of Group 3 also followed the same enrollment practice as Groups 1 and 2. Group 1 received 1 dose of the 5 ug, Group 2 received 1 dose of the 10 ug and Group 3 received 1 dose of the 20 ug.

All subjects were monitored for at least 60 minutes after each study injection. Solicited and unsolicited adverse events (AEs) were recorded for 7 days and 28 days respectively following each study injection. Serious adverse events (SAEs), including suspected and unexpected serious reaction (SUSAR), adverse events of special interest (AESIs), and medically attended adverse events (MAAEs) were recorded for the entire duration of the study. Clinical safety laboratory evaluations were performed at screening, and 7 days post each vaccination. Immune responses (antibody and cellular) were assessed at baseline and periodically after study injections. All subjects were followed up for approximately 180 days. A total of five phlebotomies were conducted during 6 clinical visits.

### Outcomes

The safety endpoints were:

1. Number and percentage of subjects with solicited local adverse events (AEs) 7 days after each vaccination.
2. Number and percentage of subjects with solicited systemic AEs 7 days after each vaccination.
3. Number and percentage of subjects with unsolicited AEs 28 days after each vaccination.
4. Number and percentage of subjects with serious adverse events (SAEs) including SUSARs throughout the study.
5. Number and percentage of subjects with Medically Attended AEs (MAAEs) throughout the study.
6. Number and percentage of subjects with AEs of special interest (AESIs) throughout the study.
7. Changes in safety laboratory parameters from baseline by FDA toxicity grading scale.

The immunogenicity endpoints were:

1. Geometric mean titer (GMT) of neutralizing antibody against wild original SARS-CoV-2 virus, pseudotype or wildtype variants of SARS-CoV-2 of concerns at baseline and pre-defined post-vaccination time points.
2. Seroconversion rate of neutralizing antibody (sero-conversion is defined by ≥ 4-fold increase from base-line)
3. Geometric mean fold increase of neutralizing antibody titer from baseline
4. GMT of serum IgG against SARS-CoV-2 by ELISA
5. Seroconversion rate of serum IgG (seroconversion is defined by ≥ 4-fold increase from baseline)
6. Frequency of CD4+ and CD8+ T cells expressing different cytokines by intracellular and extracellular staining using Spike protein overlapping peptide pool. (e.g., Intracellular: IFN-γ, TNF-α, IL-4, IL-17a. /Extracellular: IL-2, IL-4, IL-6, IL-10, TNF, IL-17A, IFN-γ)
7. Geometric mean titer (GMT) of neutralizing antibody against Delta (B.1.617.2) and Omicron (B.1.1.529) variants of SARS-CoV-2 of concerns at baseline and pre-defined post-vaccination time points.

## Statistical Analysis

The sample sizes for each of the study group and for the overall study were selected based on the planned descriptive analyses of safety and immunogenicity data rather than on formal statistical hypothesis testing. The sample size for this study was selected to provide a preliminary assessment of safety, tolerability and immunogenicity of PIKA covid-19 vaccine. The sample size was adequate for initial review of frequent adverse events rather than rare AEs. However, the probability of observing one or more AEs given various true event rates were presented in Table. If the true AE rate was 5%, given the sample size of 15 subjects in each dose group, the chance of observing at least one AE was 79.4%; with 45 subjects across three dose groups per Arm, the chance of observing of at least one AE was 90.1%.

This interim analysis was to provide information to guide the design of next studies. It was performed for Arm A and Arm B1 based on cumulative immunogenicity and safety data through day 35 for the subjects.

Safety analysis population included all subject who received one dose of vaccine. The immunogenicity analysis population will include per-protocol population (PP) and modified intent- to treat population (mITT). PP consisted of all enrolled subjects who received all scheduled doses of PIKA COIVD-19 vaccine and had no major protocol deviation. mITT population would consist of all enrolled subjects who received at least one dose of PIKA COVID-19 vaccine and contributed both pre- and at least one post-vaccination blood samples for immunogenicity testing for which valid results were reported.

## Results

This interim analysis report presented the study population results pertaining to study participants in Arm A and Arm B1 who completed Day 35 for 2 doses in Arm A and Day 28 for a single booster dose in Arm B1.

As outlined in figure 1, a total of 90 participants were equally randomized to the three doses of each study arm (A and B1), 45 subjects in Arm A and 45 subjects in Arm B1. All randomized participants according to dose and treatment constituted the safety analysis set in each arm.

**Figure 1.**
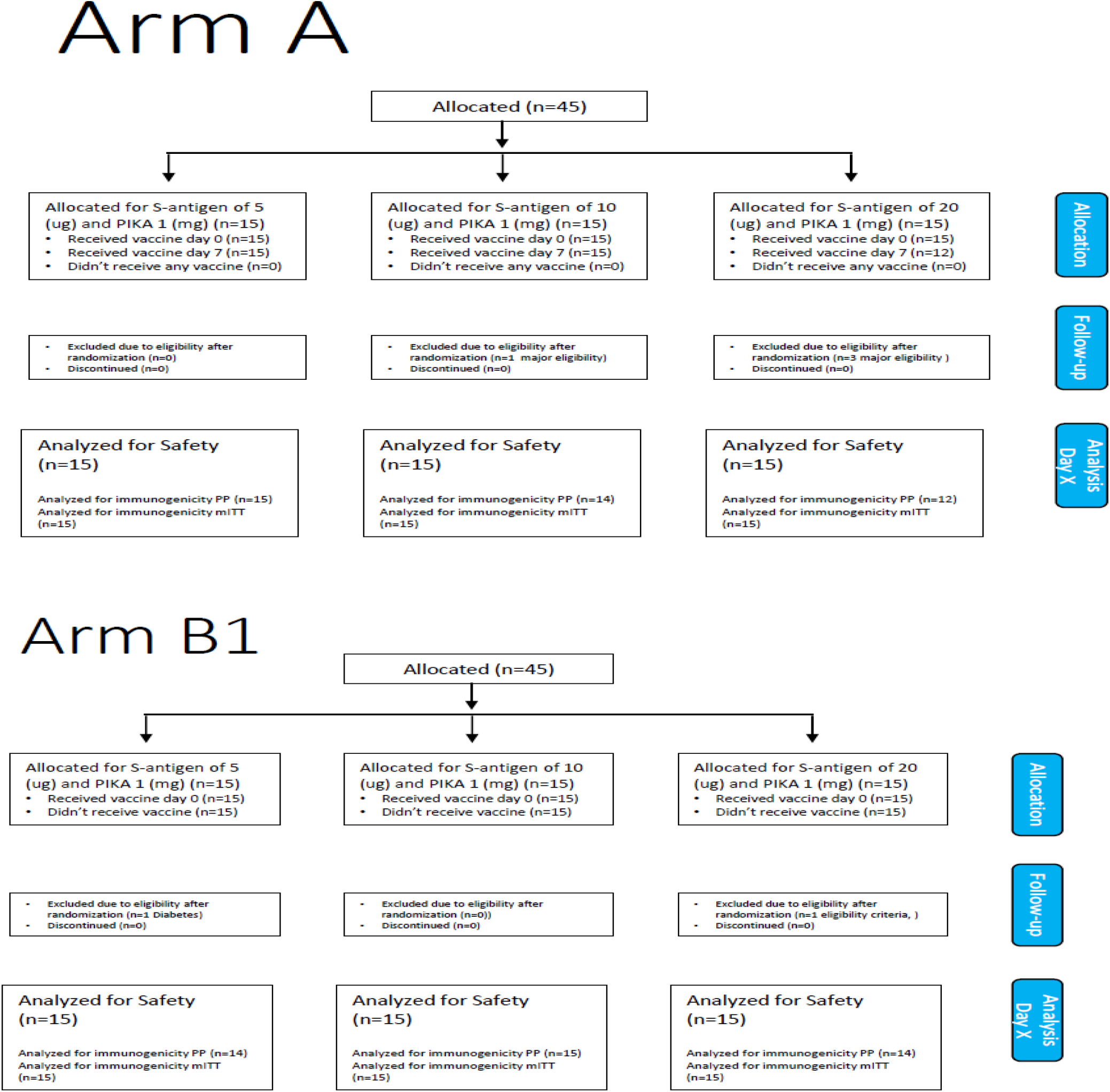
Disposition of Participants.

In arm A, a total of 45 subjects who had never received any Covid 19 vaccination or have had Covid 19 infection for > 6 months prior to enrolment were enrolled in this cohort and were assigned to 3 dose escalation groups, the 5 ug, 10 ug and the 20 ug. Each subject received the PIKA COVID 19 vaccine on Day 0 and Day 7 via intramuscular injection. Group 1 received 2 doses of the 5 ug, Group 2 received 2 doses of the 10 ug and Group 3 received 2 doses of the 20 ug.

In arm B1, a total of 45 subjects who had completed their primary series of Covid 19 vaccination with an Inactivated Covid 19 vaccine were enrolled in this cohort and were assigned to the 3 dose escalation groups, the 5 ug, 10 ug and the 20 ug. Each subject received 1 dose of the PIKA COVID 19 vaccine. Group 1 received 1 dose of the 5 ug, Group 2 received 1 dose of the 10 ug and Group 3 received 1 dose of the 20 ug.

A total of 6 participants were excluded from the Per Protocol Analysis Set (PPS) for the immunogenicity analysis. In Arm A, there were 4 participants excluded from PPS, 1 from the 10 ug and 3 from the 20 ug groups. In Arm B1, there were 2 participants excluded, 1 each from the 5 ug and 20 ug groups. These exclusions were due to positive results of SARS-CoV-2 RT-PCR which were delayed and participants were randomized using Rapid Antigen Kit which showed negative results. 2 of the exclusions were due to ineligibility criteria where participants were confirmed to have medical history of uncontrolled diabetes mellitus and Hepatitis C infection.

## Results

This interim analysis report presented the study population results pertaining to study participants in Arm A and Arm B1 who completed Day 35 for 2 doses in Arm A and Day 28 for a single booster dose in Arm B1.

As outlined in figure 1, a total of 90 participants were equally randomized to the three doses of each study arm (A and B1), 45 subjects in Arm A and 45 subjects in Arm B1. All randomized participants according to dose and treatment constituted the safety analysis set in each arm.

In arm A, a total of 45 subjects who had never received any Covid 19 vaccination or have had Covid 19 infection for > 6 months prior to enrolment were enrolled in this cohort and were assigned to 3 dose escalation groups, the 5 ug, 10 ug and the 20 ug. Each subject received the PIKA COVID 19 vaccine on Day 0 and Day 7 via intramuscular injection. Group 1 received 2 doses of the 5 ug, Group 2 received 2 doses of the 10 ug and Group 3 received 2 doses of the 20 ug.

In arm B1, a total of 45 subjects who had completed their primary series of Covid 19 vaccination with an Inactivated Covid 19 vaccine were enrolled in this cohort and were assigned to the 3 dose escalation groups, the 5 ug, 10 ug and the 20 ug. Each subject received 1 dose of the PIKA COVID 19 vaccine. Group 1 received 1 dose of the 5 ug, Group 2 received 1 dose of the 10 ug and Group 3 received 1 dose of the 20 ug.

A total of 6 participants were excluded from the Per Protocol Analysis Set (PPS) for the immunogenicity analysis. In Arm A, there were 4 participants excluded from PPS, 1 from the 10 ug and 3 from the 20 ug groups. In Arm B1, there were 2 participants excluded, 1 each from the 5 ug and 20 ug groups. These exclusions were due to positive results of SARS-CoV-2 RT-PCR which were delayed and participants were randomized using Rapid Antigen Kit which showed negative results. 2 of the exclusions were due to ineligibility criteria where participants were confirmed to have medical history of uncontrolled diabetes mellitus and Hepatitis C infection.

### Demographics

The average age of participants in Arm A and in Arm B1 were 33.6 years and 31.1 years respectively. The majority of participants in both arms were males (78% in Arm A and 88.4% in arm B1). In Arm A, almost a third of participants were Asian, another third of the participants were African/Black and a quarter of them were white/Caucasian or Arab (table 1). In Arm B, almost half of participants were Asian, 21% were Black/African and 28% were white/Caucasian or Arab (table 2). Randomization of 45 participants to the three doses in each study Arm did not produce well balanced groups in terms of the demographics due to the small sample size of the study and not using stratification in the randomization.

**Table 1.** Demographics of Arm A

| Variables |  | ARM A |  |  |  |  |  | OVERALL ARM A |  |
| --- | --- | --- | --- | --- | --- | --- | --- | --- | --- |
| | | 5 μg | | 10 μg | | 20 μg | | | |
| Age | Mean ± sd (range) | 31.7±9.5 | (21-48) | 33.0±6.5 | (23-46) | 36.7±5.5 | (26-43) | 33.6±7.6 | (21-48) |
|  | Median (q1-q3) | 31 | (23-39) | 32 | (28-36) | 37 | (33.5-41.5) | 33 | (27-40) |
| Gender |  | n | % | n | % | n | % | n | % |
|  | Female | 6 | 40.0% | 1 | 7.1% | 2 | 16.7% | 9 | 22.0% |
|  | Male | 9 | 60.0% | 13 | 92.9% | 10 | 83.3% | 32 | 78.0% |
| RACE | Asian | 8 | 53.3% | 3 | 21.4% | 3 | 25.0% | 14 | 34.1% |
|  | African/Black | 3 | 20.0% | 7 | 50.0% | 4 | 33.3% | 14 | 34.1% |
|  | White/Caucasian/Arab | 2 | 13.3% | 3 | 21.4% | 5 | 41.7% | 10 | 24.4% |
|  | Other: Pakistani/Indian/Persian | 2 | 13.3% | 1 | 7.1% | 0 | 0.0% | 3 | 7.3% |

**Table 2.** Demographics of Arm B

| Variables |  | ARM B1 |  |  |  |  |  | OVERALL ARM B1 |  |
| --- | --- | --- | --- | --- | --- | --- | --- | --- | --- |
| | | 5 μg | | 10 μg | | 20 μg | | | |
| Age | Mean ± sd (range) | 33.0±7.3 | (22-47) | 28.3±6.5 | (21-42) | 31.4±8.6 | (21-50) | 31.1±7.5 | (21-50) |
|  | median (q1-q3) | 33.5 | (28-35) | 26 | (22-33) | 30.5 | (24-36) | 31 | (25-35) |
| Gender |  | n | % | n | % | n | % | n | % |
|  | Female | 2 | 14.3% | 2 | 13.3% | 1 | 7.1% | 5 | 11.6% |
|  | Male | 12 | 85.7% | 13 | 86.7% | 13 | 92.9% | 38 | 88.4% |
| RACE | Asian | 7 | 50.0% | 8 | 53.3% | 6 | 42.9% | 21 | 48.8% |
|  | African/Black | 2 | 14.3% | 4 | 26.7% | 3 | 21.4% | 9 | 20.9% |
|  | White/Caucasian/Arab | 5 | 35.7% | 3 | 20.0% | 4 | 28.6% | 12 | 27.9% |
|  | Other: Pakistani/Indian/Persian | 0 | 0.0% | 0 | 0.0% | 1 | 7.1% | 1 | 2.3% |

### Safety

For Arm A, In each dose, 9 (60%) out of the 15 participants reported solicited adverse events after the first vaccination. However, after the second vaccination, the number of solicited adverse events decreased to 4 (26.7%), and 5(33.3%) among the 15 participants in each of doses 5μg and 10μg respectively. For the 12 participants who received the second vaccine in the high dose group, 5 (41.7%) reported any solicited adverse events. Most of the solicited events reported were local (mainly pain/tenderness) with few systemic (mainly fever and headaches). The majority of participants reported unsolicited events after the first vaccination (66.7%, 60% and 53.3% for doses 5, 10 and 20μg respectively). Most of these were results of laboratory investigations (including hematology/hepatobiliary/renal or urine analysis) followed by respiratory, thoracic and mediastinal disorders. These number decreased post second vaccination from Arm A.

There were 4 SAEs reported, 4 participants, 2 (13.3%) in the 5μg dose and 2 (13.3%) in the 20μg dose were diagnosed with Covid19 infection. These SAEs were not related to the study vaccine and not expected.

Two participants experienced medically attended Adverse events. One (6.7%) in each of the 5μg dose and 10μg dose groups. Descriptions of those are included in this report. The vast majority of participants reported mild AEs (at least 80% in each dose group) and majority of the participants reported both unexpected and expected events, and also definitely related/possibility related and not related/unlikely not related events.

The most common solicited local reaction reported in all dose group in Arm A is injection site pain/tenderness. After the first vaccination, a total of 21 participants reported injection site pain/tenderness; 6 (40.0%) from the 5 ug, 7 (46.7%) from the 10 ug and 8 (53.3%) from the 20 ug. Other solicited local reactions reported post first vaccination were erythema/redness on injection site (one each from the 10 ug and 20 ug). After the second vaccination, only 8 participants reported injection site pain/tenderness; 1 (6.7%) from the 5 ug, 2 (13.3%) from the 10 ug and 5 (41.7%) from the 20 ug. Erythema/redness on injection site was reported only from 1 participant (6.7%) from the 10 ug and there were 2 cases of swelling/induration on injection site reported from 1 participant each from the 5 ug and 10 ug dose group. For the solicited systemic adverse events, fever or pyrexia was the most common solicited systemic reaction. After the first vaccination, fever was reported by 1 (6.7%) participant from the 5 ug, 1 (6.7%) from the 10 ug and 2 (13.3%) from the20 ug dose group.

After the second vaccination, only 4 participants reported fever/pyrexia, 2 (13.3%) from the 5 ug and 2 (13.3%) from the 10 ug dose group. Other systemic solicited adverse reactions reported were myalgia, headache, fatigue/malaise.

All solicited local and systemic AEs were mild to moderate and resolved within a few days with or without medication. Unsolicited adverse events reported up to 35 days after the 1st vaccination (or 28 days after the 2nd vaccination) were unremarkable. There was no AESI reported from Arm A.

For Arm B1, 33.3%, 60.0% and 46.7% of the participants in each of the 5, 10 and 20μg dose groups respectively reported solicited adverse events after the vaccine. Most of the solicited events reported were local (only pain/tenderness) with few systemic (mainly fever and headaches). 66.7%, 33.3% and 33.3% of the participants in doses 5, 10 and 20μg groups respectively reported unsolicited events after vaccine. Most of those were investigations (including hematology/hepatobiliary/Renal or Urine tract infection urine analysis). No SAE, AESI and no medically attended adverse events were reported in Arm B1.

The majority of participants reported mild AEs (at least 60% in each dose group) and between 40 and 60% of the participants reported both unexpected and expected events, and majority reported AEs with relationship to vaccine as definitely related/possibility related and AEs with relationship to vaccine as not related/unlikely not related events.

In Arm B1, the most common and only reported solicited local reaction was still pain/tenderness on injection site which was reported from a total of 16 participants, 3 (20.0%) from the 5 ug, 8 (53.3%) from the 10 ug and 5 (33.3%) from the 20 ug dose groups. Fever was the most common solicited systemic reaction being reported from 1 (6.7%) from the 5 ug and 2 (13.3%) from the 10 ug dose groups. Headache was reported from 1 (6.7%) from the 10 ug and 4 (26.7%) from the 20 ug dose groups. Other solicited systemic reactions reported were 1 case each of myalgia and fatigue/malaise.

All solicited local and systemic AEs were mild to moderate and resolved within a few days. Unsolicited adverse events reported up to 28 days post vaccination were unremarkable.

### Screening laboratory tests

Screening laboratory tests (including hematology, biochemistry, coagulation, and urinalysis) were performed during the screening process and were repeated on Day 7 and Day 14 in both Arm groups. Results from these laboratory tests during screening served as study-entry baseline values. Abnormal results and findings that make the subject ineligible were discussed with the subject and the subjects were referred for follow-up care with their healthcare provider if necessary.

In arm A, laboratory tests assessment revealed no hematological abnormality of clinical significance, one urine RBC abnormality of clinical significance in the 10μg dose group at day 14, and three participants with glucose presence in urine: one at screening in the 10μg dose group, one at day 7 in the 5μg dose group and one at day 7 in the 20μg dose group. Moreover, there were no abnormalities of clinical significance in serum biochemistry laboratory results and one abnormality of clinical significance in Coagulation test PT at day 14 in the 20μg dose group. All significant laboratory result findings were reported as AEs and were followed up until resolution.

In arm B1, laboratory tests assessment revealed no hematological abnormality of clinical significance, one urine WBC abnormality of clinical significance in the 10μg dose group at day 14, and five incidences with glucose presence in urine: one at screening, one at day 7 and one at day 14 in the 5μg dose group, and one at screening and one at day 14 in the 20μg dose group. Moreover, there were 4 abnormalities of unknown clinical significance: 2 in serum AST at days 7 and day14 of the 10μg dose group and also 2 in ALT at the same days and same dose group. Finally, there were no abnormalities of clinical significance in coagulation test in all dose groups.

### Immunogenicity

Figure 2 presented the comparison data for GMTs of neutralizing antibodies against the wild type virus, delta and omicron for all 3 dose groups of 5 μg, 10 μg and 20 μg in Arm A.

**Figure 2.**
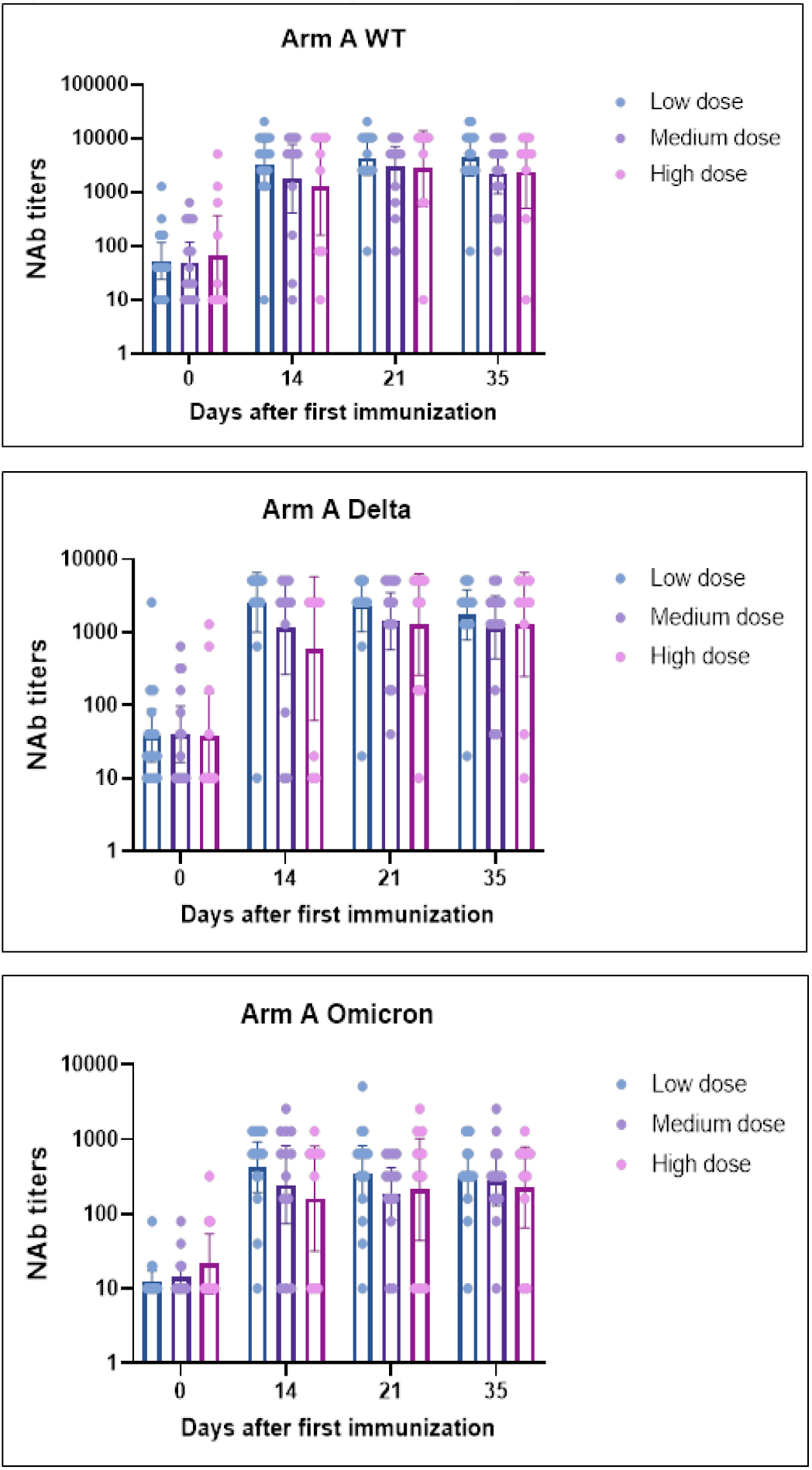
GMTs of neutralizing antibodies against 3 variants (Arm A)

There was significant increase in GMTs of neutralizing antibodies for all three variants (Wild type, Delta and Omicron) from Day 0 (before vaccination) to Day 14 in all three dose groups. For the Wild type virus, GMT was shown to still increased on Day 21 and slightly decreased by Day 35. For the other two variants (Delta and Omicron), there was a decrease from Day 14 at both Days 21 and 35. However, in all three dose groups and for all three variants the change from baseline Day 0 was substantial.

On Day 35 or 28 days post the second vaccination, GMTs of neutralizing antibody against the wild type virus was 4413.3 (1976.8, 9772.4) for the 5 μg, 2206.7 (931.6, 5226.9) for the 10 μg and 2398.8 (501.2, 11220.2) for the 20μg dose group at 95% CI. GMTs of neutralizing antibody against the Delta variants was 1722.8 (777.2, 3818.8) for the 5 μg, 1148.2 (430.3, 3123.2) for the 10 μg and 1288.2 (251.2, 6606.9) for the 20μg dose group. GMTs of neutralizing antibody against the Omicron variant was 323.6 (154.9, 660.7) for the 5 μg, 275.8 (129.5, 587.3) for the 10 μg and 223.9 (66.1, 776.2) for the 20μg dose group at 95% CI.

Figure 3 presented the comparison data for seroconversion rates of neutralizing antibodies against the wild type virus, delta and omicron for all 3 dose groups of 5 μg, 10 μg and 20 μg in Arm A.

**Figure 3.**
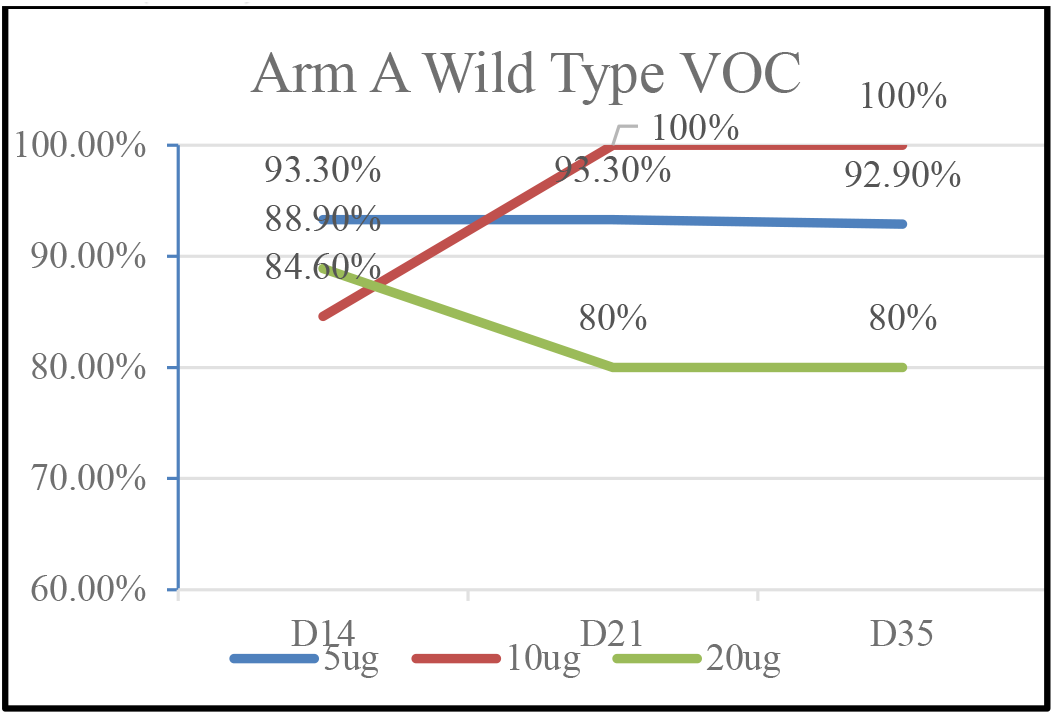
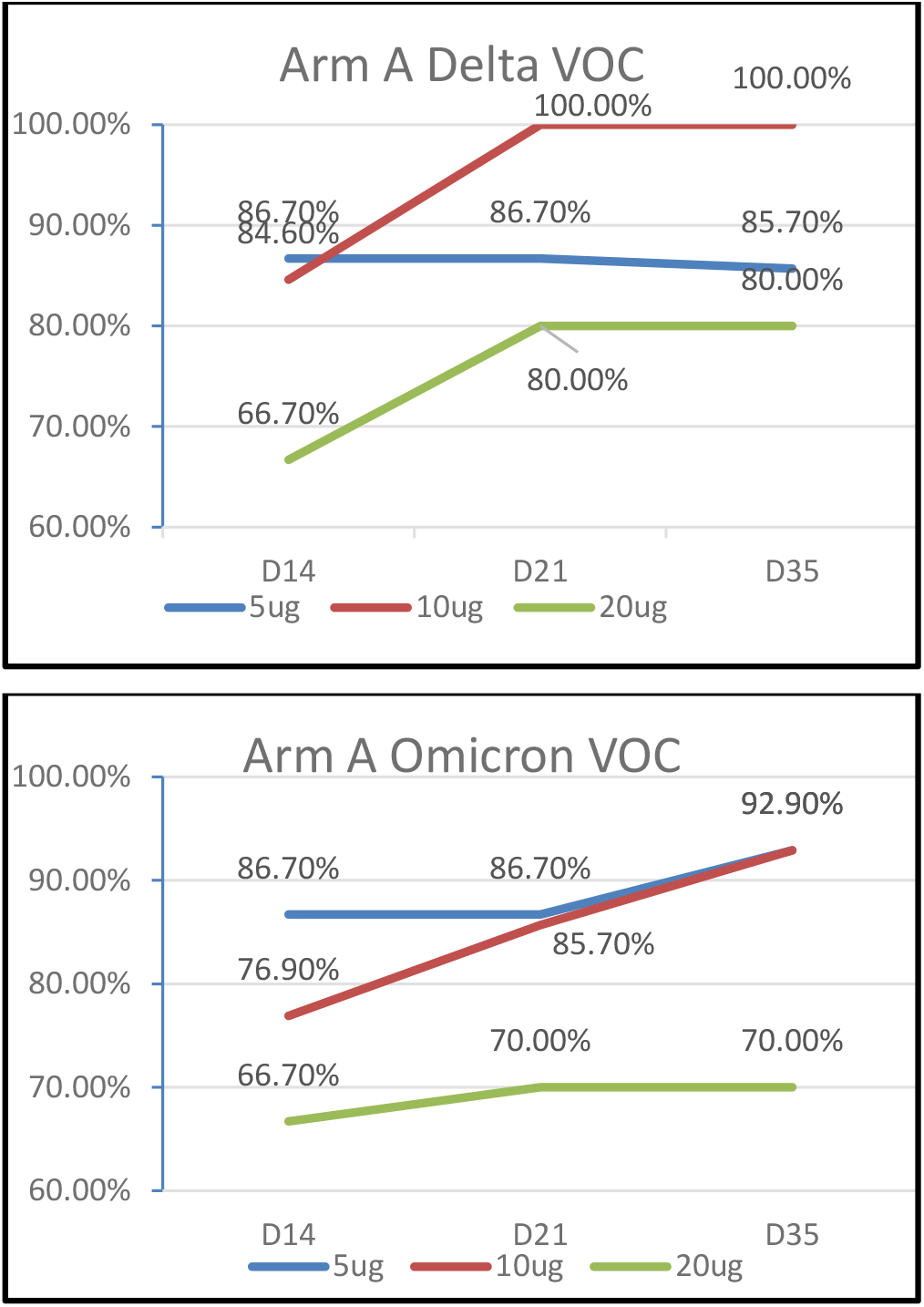
Seroconversion rates of neutralizing antibodies against 3 variants (Arm A)

Generally, at Days 14, 21 and 35 and for the Wild type, Delta and Omicron variants, the seroconversion rates were very high in dose groups 5 μg (ranging from 85.7% to 93.3%) and in dose group 10 μg (ranging from 76.9% to 100%). These rates were slightly lower for the 20μg group (ranging from 66.7% to 88.9%) possibly since this later group had larger baseline values.

At Day 35 or 28 days post the second vaccination, seroconversion rate of neutralizing antibody against the wild type virus was 92.9% (71.2, 99.2) for the 5 μg, 100% (76.8, 100) for the 10 μg and 80.0% (49.7, 95.6) for the 20 μg at 95% CI. Seroconversion rate of neutralizing antibody against the Delta variant was 85.7% (61.5, 96.9) for the 5 μg, 100% (76.8, 100) for the 10 μg and 80.0% (49.7, 95.6) for the 20 μg. Lastly, the seroconversion rate of neutralizing antibody against the Omicron variant was 92.9% (71.2, 99.2) for the 5 μg, 92.9% (71.2, 99.2) for the 10 μg and 70.0% (39.4, 90.7) for the 20 μg at 95% CI.

Figure 4 presented the comparison data for GMTs of neutralizing antibodies against the wild type virus, delta and omicron for all 3 dose groups of 5 μg, 10 μg and 20 μg in Arm B1.

**Figure 4.**
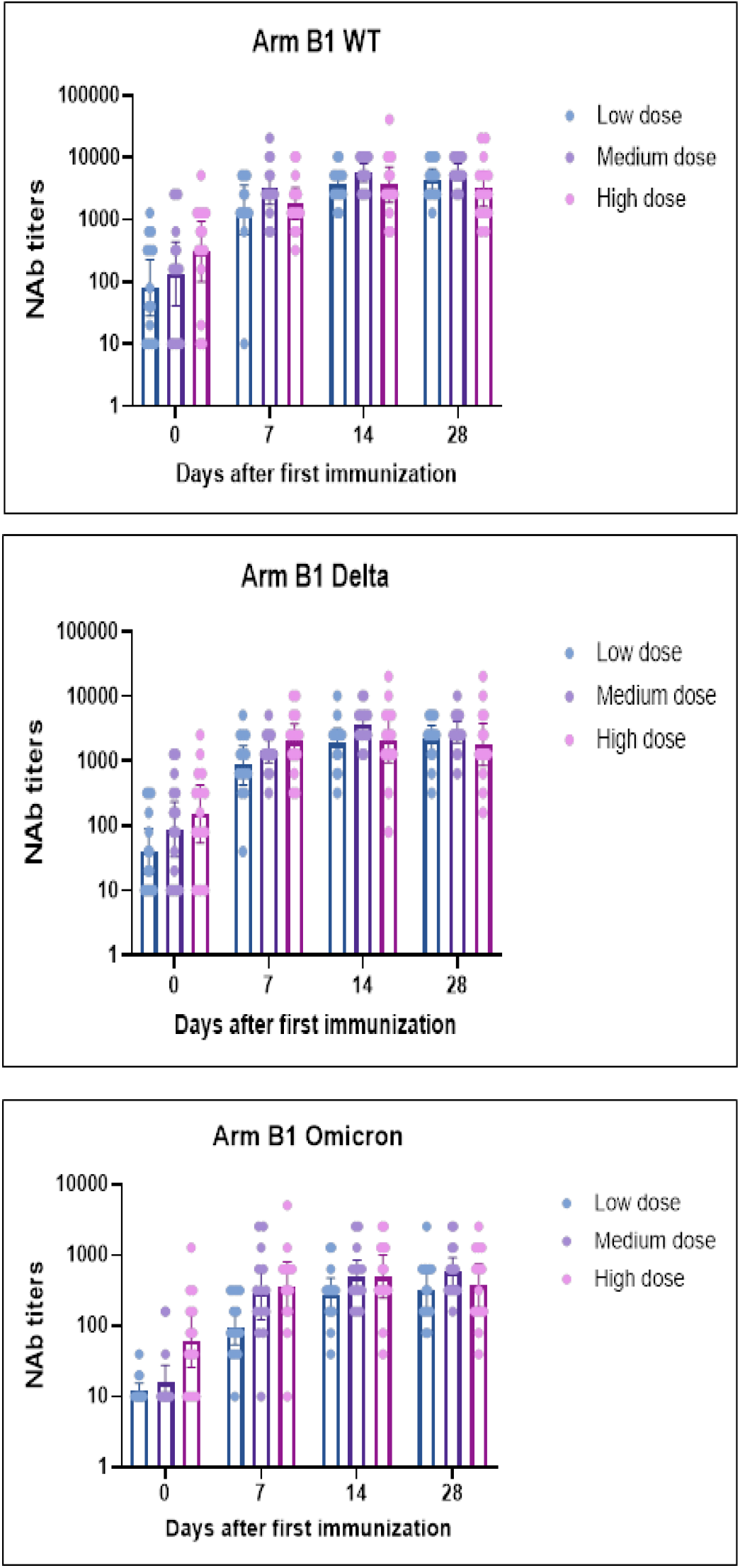
GMTs of neutralizing antibodies against 3 variants (Arm B1)

Generally, there was a significant increase in GMTs of neutralizing antibodies for all three variants (wild type, delta and omicron) from Day 0 (before vaccination) to Day 28 in all three dose groups with few exceptions of a small decrease at Day 28 compared to Day 14 especially in the 20μg dose group. However, in all three dose groups and for all three variants the change from baseline day 0 is substantial.

At Day 28 or 28 days post the single booster vaccination, GMTs of neutralizing antibody against the wild type virus was 4413.3 (2987.5, 6519.6) for the 5 μg, 5881.3(4368.6, 7917.8) for the 10 μg and 3120.7 (1620.5, 6009.8) for the 20μg dose group at 95% CI. GMTs of neutralizing antibody against the delta variants was 2206.7 (1371.5, 3550.4) for the 5 μg, 2807.9 (1919.9, 4106.6) for the 10 μg and 1810.2 (856.2, 3827.3) for the 20μg dose group. GMTs of neutralizing antibody against the omicron variant was 320.0 (181.7, 563.6) for the 5 μg, 583.5 (369.9, 920.4) for the 10 μg and 371.2 (183.4, 751.4) for the 20μg dose group at 95% CI.

Figure 5 presented the comparison data for seroconversion rate of neutralizing antibodies against the wild type virus, delta and omicron for all 3 dose groups of 5 μg, 10 μg and 20 μg in Arm B1.

**Figure 5.**
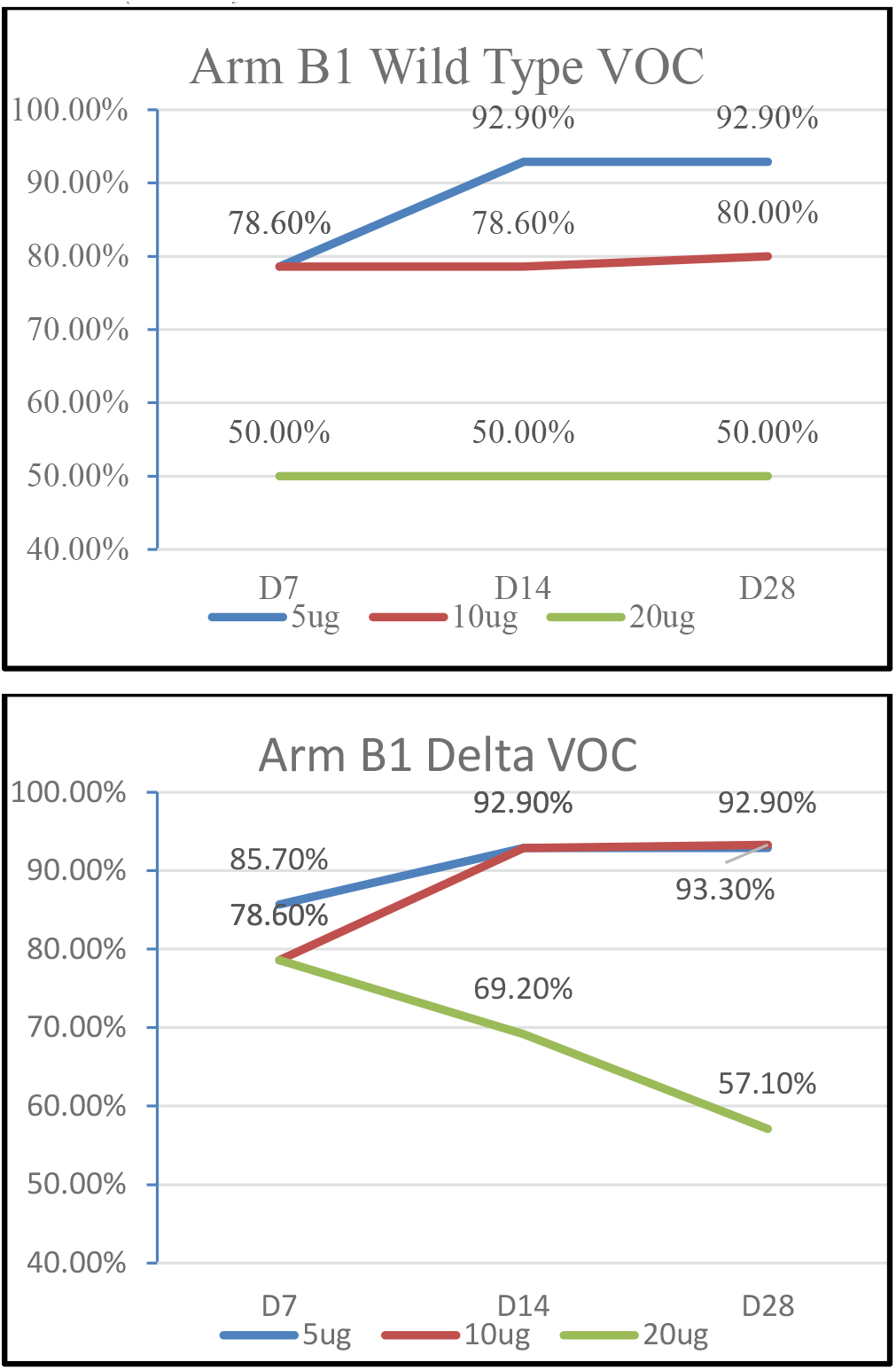
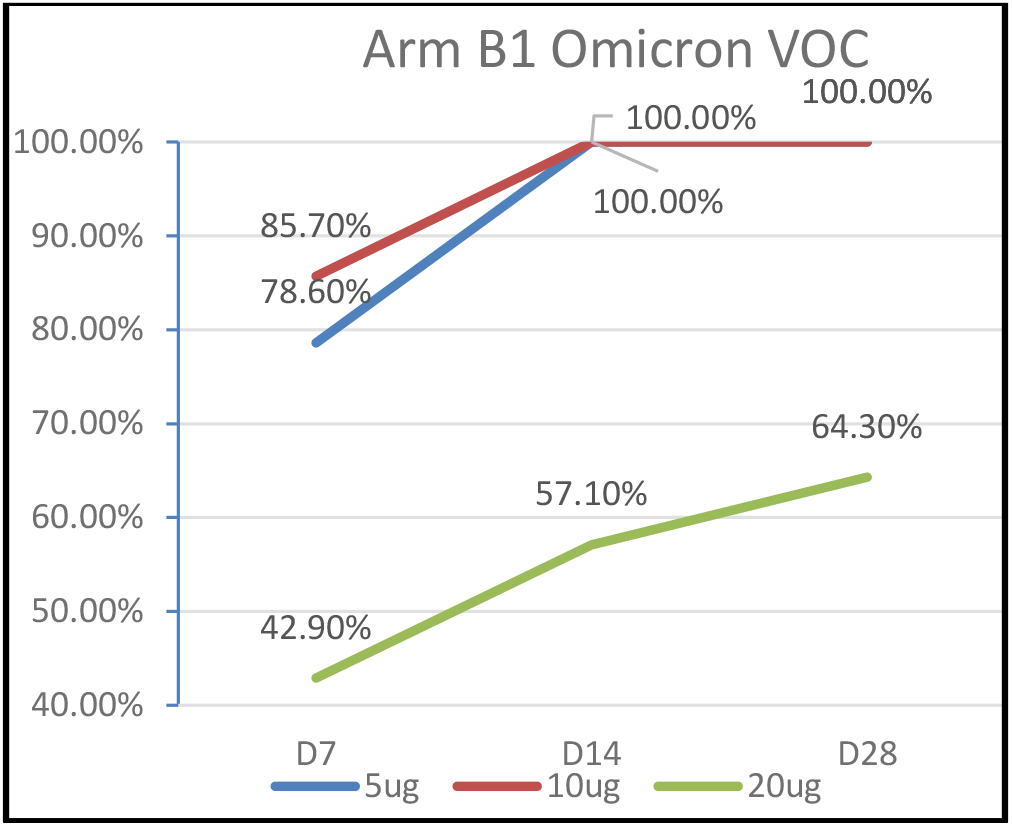
Seroconversion rates of neutralizing antibodies against 3 variants (Arm B1)

Generally, at Day 28 and for all variants, the seroconversion rates were very high in dose groups 5 μg (ranging from 92.9% to 100%) and in dose group 10 μg (ranging from 80% to 100%). The rates were lower for the 20μg group (ranging from 50% to 64.3%) possibly since this later group had larger baseline values.

At Day 28 or 28 days post the single booster vaccination, sero-conversion rate of neutralizing antibody against the wild type virus was 92.9% (71.2, 99.2) for the 5 μg, 80.0% (55.6, 94.0) for the 10 μg and 50.0% (25.9, 74.1) for the 20 μg at 95% CI. Seroconversion rate of neutralizing antibody against the delta variant was 92.9% (71.2, 99.2) for the 5 μg, 93.3% (72.8, 99.3) for the 10 μg and 57.1% (31.9, 79.7) for the 20 μg. Lastly, the seroconversion rate of neutralizing antibody against the omicron variant was 100% (76.8, 100.0) for the 5 μg, 100% (78.2, 100.0) for the 10 μg and 64.3% (38.5, 84.9) for the 20 μg at 95% CI.

## Discussion and Conclusion

This was the first-in-human study for Yisheng Biopharma’s PIKA Covid 19 vaccine to assess the safety, tolerability and immunogenicity of the PIKA Covid 19 vaccine in naïve subjects or subjects who had previous Covid 19 infection for at least 6 months prior to study participation (Arm A) and as a booster dose in subjects who had completed their primary series of Covid 19 vaccination with an inactivated Covid 19 vaccine.

Safety had been evaluated for the PIKA Covid 19 vaccine in both Arm A and Arm B1 in terms of immediate reactions for the first 60 minutes after vaccine administration, solicited and unsolicited adverse events (AEs) for 7 days and 28 days, respectively following study vaccine administration were also collected. Serious adverse events (SAEs), including suspected and unexpected serious reaction (SUSAR), adverse events of special interest (AESIs), and medically attended adverse events (MAAEs) were likewise recorded until Day 35 for Arm A and Day 28 for Arm B1 for this interim report. Clinical safety laboratory evaluation including urinalysis, hematology, biochemistry and coagulation were performed at baseline and repeated on Days 7 and 14 to check whether the study vaccine would produce any significant safety concern on these parameters.

Safety profile was satisfactory. There were no significant immediate reactogenicity reported in both Arm A and Arm B1. After 7 days post vaccination, the most common solicited local reaction reported in both groups was injection site pain/tenderness and fever/pyrexia was the most common solicited systemic reaction reported. All these reactions were noted to have decreased after the second vaccination in Arm A and were resolved in a few days.

In Arm A, PIKA Covid 19 vaccine was administered in a 2-dose regimen on Day 0 and Day 7 as a primary series for Covid 19 vaccination. There was significant increase in GMTs of neutralizing antibodies against the wild type virus, Delta and Omicron in all dose groups of 5 μg, 10 μg and 20 μg after the second dose on Day 14. Although there was slight decrease in the GMTs at Day 35 or 28 days post the second dose, the level of immune response was still sustained at a very high level. Seroconversion rates at Days 14, 21 and 35 and for the Wild type, Delta and Omicron variants were very high in dose groups 5 μg (ranging from 85.7% to 93.3%) and in dose group 10 μg (ranging from 76.9% to 100%). The rates were slightly lower for the 20μg group (ranging from 66.7% to 88.9%).

In Arm B1, PIKA Covid 19 vaccine was administered as a single booster dose on Day 0 to participants who have completed their primary series of inactivated Covid 19 vaccination. Likewise, the GMTs of neutralizing antibodies against the wild type virus, Delta and Omicron in all dose groups of 5 μg, 10 μg and 20 μg showed significant increase by Day 7. In the 5μg group, the GMTs continued to increase up to Day 28 for the wild type virus, Delta and Omicron. In the 10μg group, GMTs of neutralizing antibodies against the wild type virus and Omicron continued to rise up to Day 28. Although with significant increase at Day 7 and Day 14, GMTs of neutralizing antibodies against all three variants were noted to be slightly decreasing in the 20μg group. Seroconversion rates at Day 28 and for all variants were very high in dose groups 5 μg (ranging from 92.9% to 100%) and in dose group 10 μg (ranging from 80% to 100%). The rates were lower for the 20μg group (ranging from 50% to 64.3%) possibly since this later group had larger baseline values.

In conclusion, the findings demonstrated that the PIKA Covid 19 vaccine is safe, well tolerated, immunogenic and can be used as a primary vaccination or as a booster dose in participants who had completed an inactivated Covid 19 vaccination primary series. A comparison of the immune responses presented in this interim analysis showed that geometric mean titer (GMTs) of neutralizing antibody against wild type of SARS-CoV-2 virus, Delta and Omicron of the 5 μg group was higher than the 10 μg and 20 μg, therefore the 5 μg was selected as the recommended dose for the Phase II and III clinical development of the PIKA Covid 19 vaccine.

## Data Availability

All data produced in the present study are available upon reasonable request to the authors

## Contributors

All authors had full access to all data in the studies and had final responsibility for the decision to submit for publications. Lai Hock Tan drafted the manuscript. Zenaida Reynoso Mojares had oversight of data collection and cleaning and critically reviewed the manuscript.

## Acknowledgements

The authors would like to thank all the participants who volunteered for this study.

